# HEART RATE AND LEFT VENTRICULAR REMODELING AFTER REPAIRED COARCTATION OF THE AORTA: A CROSS-SECTIONAL STUDY

**DOI:** 10.64898/2026.02.16.26346437

**Authors:** Marina Vaccari, Laura E. Maldonado, Claudio G. Moros, Angela Sardella, Miriam Romo, César A. Romero, the ARECO study group

## Abstract

**Background:** Patients with repaired coarctation of the aorta (CoAo) remain at risk for left ventricular hypertrophy (LVH) even in the absence of hypertension. Alterations in wave reflection and the timing of reflected pressure waves may contribute to ventricular remodeling beyond pressure load alone.

**Methods:** We performed a cross-sectional analysis of patients with repaired CoAo. Office and ambulatory blood pressure (ABPM), non-invasive central hemodynamics, and echocardiographic indices of left ventricular structure were assessed. Linear and multivariable regression models evaluated associations with posterior wall thickness (PWTd) and interventricular septal thickness (IVSTd). Computational simulations were conducted to examine the impact of heart rate on ventricular remodeling.

**Results:** Fifty-seven patients (median post-repair follow-up 11 years) were included. LVH prevalence was 15.2% (95% CI: 4.8–25.6). Although 42% met criteria for hypertension based on ABPM, no patients exhibited elevated central blood pressure. Adjusted augmentation index (AIX@75) was inversely associated with PWTd and remained independently associated after multivariable adjustment (R^2^ = 0.40, p < 0.01). Replacing AIX@75 by heart rate improved model performance (R^2^ = 0.44), with lower heart rate independently associated with greater PWTd. Simulation modeling showed that a 10% increase in heart rate reduced mean PWTd and decreased posterior wall hypertrophy prevalence from 30.9% to 2.4% (OR =0.10; 95% CI: 0.01–0.44).

**Conclusions:** Ventricular remodeling occurs despite normal central blood pressure in CoAo. A lower heart rate associates with increased ventricular mass. Heart rate–mediated modulation of wave reflection timing represents a potential mechanistic and therapeutic target.

## Introduction

Coarctation of the aorta (CoAo) is a common congenital cardiopathy, occurring in approximately 3.7 per 10,000 live births and representing a frequent cause of secondary hypertension in children.(1,2) Despite early repair, patients with CoAo continue to exhibit a high incidence of new-onset hypertension during follow-up and a disproportionately elevated burden of cardiovascular disease across adulthood.(3) Even among individuals who remain normotensive after repair, long-term survival is reduced, and early cardiovascular events remain common.(4,5)

Left ventricular hypertrophy (LVH) typically regresses after correction of CoAo. However, a substantial proportion of patients, despite adequate blood pressure control, develop left ventricular (LV) remodeling within a few years of repair.(6) The mechanisms driving this maladaptive remodeling remain incompletely understood. LV geometric changes, including increased thickness of the posterior wall and interventricular septum, have been reported even when values fall within age- and body size–adjusted reference ranges.(6) These subtle structural abnormalities have been associated with higher cardiovascular risk, suggesting persistent hemodynamic stress despite normalized blood pressure.(4,7)

Multiple factors influence LV remodeling in repaired CoAo(8,9) A growing body of evidence indicates that in addition to blood pressure and body size, residual vascular abnormalities, such as aortic stiffness, fibrotic scarring at the repair site, and altered arterial mechanics may play a key role in LV remodeling.(10,11) These changes can amplify LV afterload by promoting the early return of reflected pressure waves.(8) The timing of this reflected wave is clinically relevant. When the backward wave arrives during early systole, it augments systolic load at the most vulnerable phase of ventricular contraction, potentially accelerating concentric remodeling and impairing long-term ventricular function.(12)

Heart rate (HR) is a critical determinant of the reflected wave timing.(13) Lower HR prolongs the cycle, making it more likely that reflected waves return during early systole and modifying the Augmentation Index.(14) Despite these physiologic considerations, the potential contribution of HR to persistent LV remodeling in repaired CoAo has not been systematically evaluated.

The objective of this study is to analyze the determinants of LV remodeling in patients with repaired CoAo. We hypothesize that aortic stiffness and HR are independent predictors of LV remodeling, and that modulation of HR may represent a potential strategy to mitigate long-term cardiovascular risk in this population.

## Material and Methods

### Study design and population

This was a retrospective, analytical, and observational study based on a review of medical records of all patients followed at the pediatric hypertension clinic of Hospital de Niños “Dr. Ricardo Gutiérrez” (Buenos Aires, Argentina) with a diagnosis of CoAo who had undergone central hemodynamic assessment.

Inclusion criteria were: (1) patients with a diagnosis of CoAo, including mild native CoAo and patients treated by surgery or interventional catheterization; and (2) age ≥8 years, corresponding to the minimum age at which central hemodynamic assessment can be reliably performed. Exclusion criteria were: (1) recoarctation of the aorta, defined by a descending aortic gradient >30 mmHg, presence of diastolic runoff, and loss of pulsatility in the abdominal aorta; (2) associated complex congenital heart disease; (3) moderate or severe aortic valve disease (stenosis and/or regurgitation), due to superimposed ventricular volume or pressure overload.

A medical record review was performed by a single investigator. Data collected included the date and type of CoAo repair; anthropometric measurements calculated as weight, height, and body mass index (BMI = weight/height^2^); blood pressure recordings; central hemodynamics; echocardiographic parameters; and laboratory data from the most recent follow-up visit. BMI, Pulse wave velocity, and left ventricular mass (LVM) were also expressed as Z-Score to the normal distribution. A Z-value of 0 represents the normal mean value, and the normal 95% confidence interval is −2 to +2.

### Blood pressure assessment

Office blood pressure was measured using auscultatory method for pediatric populations, with an appropriately sized cuff selected in accordance with guideline recommendations.(15,16) Three blood pressure measurements were obtained in all four limbs. The mean of three measurements obtained in the right arm was used for analysis, except in cases with documented right subclavian artery involvement. Blood pressure values were expressed as percentiles according to national reference tables adjusted for age, sex, and height.(17) Office hypertension was defined as systolic and/or diastolic blood pressure ≥95th percentile in patients <16 years of age, and ≥140/90 mmHg in patients ≥16 years of age.

ABPM was performed using validated pediatric devices (Spacelabs 90210), with measurements taken every 20 minutes during the day and every 30 minutes at night. Daytime and nighttime periods were defined according to patient-reported activity. Hypertension was defined as mean daytime and/or nighttime blood pressure ≥95th percentile according to sex- and height-adjusted reference values.(15) A non-dipping pattern was defined as a nocturnal blood pressure reduction <10%. ABPM recordings were considered valid if ≥70% of measurements were successful and at least seven readings were available during the sleep period. Patients receiving antihypertensive medication and those with ambulatory hypertension were classified as hypertensive. White-coat hypertension was defined as elevated office blood pressure with normal ABPM, while masked hypertension was defined as normal office blood pressure with elevated ABPM.

### Vascular Mechanics and Echocardiography

Vascular mechanics were assessed using an oscillometric device (Mobil-O-Graph, IEM). Measurements were obtained after 5–10 minutes of rest, under conditions similar to those used for office blood pressure measurement. The device automatically evaluated signal quality and repeated measurements until three high-quality recordings were obtained. The average of the three recordings was used for analysis. Parameters analyzed included pulse wave velocity (PWV), central blood pressure (CBP), and heart rate– adjusted augmentation index (AIX@75). Arterial stiffness was defined as PWV above the 90th percentile adjusted for age and sex according to reference values.(18)

Transthoracic echocardiography was performed using a Siemens Acuson SC2000 Sequoia system with a 2.5-MHz Acuson 4V1c transducer. Echocardiographic data were obtained using two-dimensional and Doppler techniques in accordance with international pediatric echocardiography guidelines.(19) Linear measurements of the left ventricle were obtained in M-mode, guided by two-dimensional imaging in parasternal long- and short-axis views. Final values represented the mean of three cardiac cycles. LVM was calculated using the Devereux formula and indexed to height raised to the 2.7 power.(20) Interventricular septal thickness in diastole (IVSTd), posterior wall thickness in diastole (PWTd), and indexed LVM were analyzed using national pediatric reference percentiles.(21) Increased wall thickness and LVH were defined as values above the 90th percentile.

### Statistical analysis

Continuous variables were assessed for distribution using graphical inspection and the Shapiro–Wilk test. Normally distributed variables are presented as mean ± standard deviation, and non-normally distributed variables as median and interquartile range. Categorical variables are presented as absolute frequencies and percentages. Patients were stratified by LVM Z-score tertiles to evaluate clinical, vascular, and echocardiographic characteristics across strata. Group comparisons were performed using the chi-square test for categorical variables and the two-way ANOVA test for continuous variables, as appropriate. Correlations between variables were assessed using Pearson’s correlation coefficient. A two-sided p-value <0.05 was considered statistically significant.

Given that LVM is determined by posterior wall and septal thickness and is derived from a nonlinear cubic formula, PWTd and IVSTd were used as primary outcomes to identify associated variables via correlation analyses. Variables with correlation coefficients < 0.10 were included in simple and multivariable linear regression models using a stepwise approach to identify the most relevant determinants. Model selection was based on the coefficient of determination, lowest mean squared error, and parsimony. Height, age, and BMI were retained in all models. Multicollinearity and interaction were assessed using variance inflation factors and interaction terms.

To assess the impact of HR on PWTd, a counterfactual simulation was performed using coefficients from the fitted multivariable linear regression model. Starting from the analytic dataset, HR was uniformly increased by 10% (HR_sim = HR × 1.10). Predicted PWTd values under the simulated HR were generated using the regression equation: PWTd_sim = β0 + β_age·age + β_SBP24·SBP24 + β_height·height + β_BMI·BMI + β_HR·HR_sim. Observed and simulated PWTd values were compared using descriptive statistics and paired t-tests. Additional stratified paired t-tests were performed by baseline posterior wall thickness category to assess differential effects. All analyses and simulations were conducted using SAS version 9.4 (SAS Institute Inc., Cary, NC, USA).

### Ethical considerations

This report presents partial results from a research protocol approved by the Ethics Committee of the Government of the Autonomous City of Buenos Aires, Hospital de Niños “Dr. Ricardo Gutiérrez” (approval code: 16417). The study was conducted in accordance with the principles of the Declaration of Helsinki. Given the retrospective design, the Ethics Committee waived the requirement for informed consent. All data were anonymized before analysis to ensure confidentiality and the protection of personal information. This study was conducted and is reported in accordance with the Strengthening the Reporting of Observational Studies in Epidemiology (STROBE) guidelines for observational studies.(22)

## Results

All patients with a diagnosis of CoAo followed at the hypertension clinic who underwent non-invasive central hemodynamic assessment between 2018 and 2025 were screened (n = 58). Patients with recoarctation of the aorta (n=1) were excluded. The final analytic cohort included 57 patients with a median post-repair follow-up of 11 years (95% CI: 9–14). Baseline characteristics of the study population are shown in Table 1.

**Table 1.** Clinical, Hemodynamic, and Echocardiographic Characteristics by LV Mass Z-Score Tertiles.

|  | Total<br>(n=57) | LV mass<br>Tertile 1<br>(n=16) | LV mass<br>Tertile 2<br>(n=16) | LV Mass<br>Tertile 3<br>(n=16) | P |
| --- | --- | --- | --- | --- | --- |
| Age (years, IQR) | 12.4 (10-15) | 12.6 (10.5-14.5) | 12.2 (9.5-14.5) | 11.7 (9-14) | 0.67 |
| Male (n,%) | 35 (61.4%) | 12(75%) | 8(50%) | 10(62.5%) | 0.34 |
| Time from surgical repair (years) | 11.2 (9-14.8) | 10.6 (9.9-14.6) | 11.8(10.2-15) | 10(8.4-13) | 0.59 |
| Height (mts) | 139.1±40.5 | 150.7±17.1 | 150.4±18.9 | 146.6±16.9 | 0.65 |
| BMI | 21.46±3.94 | 20.24±3.42 | 20.72±3.5 | 23.8±4.2 | **0.01** |
| BMI z score | 1.22±1.42 | 0.8±1.4 | 1±1.2 | 2.2±1.5 | **0.01** |
| BSA | 1.37±0.34 | 1.39±0.28 | 1.4±0.32 | 1.42±0.3 | 0.97 |
| HR (BPM) | 77±10.9 | 75.2±10.31 | 78.92±10.5 | 77.9±12.36 | 0.64 |
| Systolic Aortic Pressure | 99.5±9.1 | 94.4±8.8 | 100.3±7.8 | 102.7±8.9 | **0.02** |
| 24 hs SBP (mmHg) | 119.4±10.9 | 115.6±10.9 | 118.4±8.9 | 123.3±13.4 | 0.19 |
| Normotension (n,%) | 34/47 (72.3%) | 11 (68.7%) | 8 (50%) | 10 (62.5%) | 0.72 |
| PWV (m/seg) | 4.6±0.3 | 4.5±0.3 | 4.7±0.4 | 4.6±0.3 | 0.23 |
| PWV z score | 0.3±1.08 | -0.1±1.4 | 0.6±1 | 0.4±0.8 | 0.13 |
| Arterial Stiffness (%) | 4/55 (7.3%) | 1 (6.25%) | 2(12.5%) | 1(6.25%) | 0.76 |
| AIX@75 | 13.04±9.1 | 12.43±9.7 | 15.25±8.4 | 12.02±9.4 | 0.57 |
| LVM (g/m<sup>2.7</sup>) | 28.9±8.8 | 20.9±4.1 | 26.5±2.6 | 39.4±5.7 | **<0.01** |
| PWTd | 6.7±1.4 | 5.9±1.2 | 6.4±1.1 | 7.9±1.2 | **<0.01** |
| IVSTd | 6.6±1.4 | 5.5±0.9 | 6.5±1 | 7.8±1.2 | **<0.01** |

The majority of patients (91.2%) had undergone corrective intervention. Surgical repair was performed in 44 patients (77.1%), while 7 patients (12.3%) underwent angioplasty with stent placement for native CoAo, and 1 patient (1.7%) was treated with angioplasty alone. Five patients (8.8%) had mild CoAo and remained under clinical follow-up without intervention.

Surgical procedures were performed between 2001 and 2014. The most frequent techniques were extended end-to-end anastomosis (31.8%), followed by simple end-to-end anastomosis (20.4%), subclavian flap repair (6.8%), and patch repair (2.3%). Nine patients (20.4%) were operated on within the first month of life and required extended aortic reconstruction. Most patients (81.8%) underwent repair within the first year of life

Regarding blood pressure status, 33 patients (58%) were normotensive and not receiving antihypertensive medication, whereas 24 patients (42%) met criteria for hypertension. Among hypertensive patients, treatment and control rates were 79% (19/24) and 58% (11/19), respectively. No cases of white-coat hypertension were identified, and only two patients (8%) exhibited masked hypertension.

Complete non-invasive central hemodynamic assessment was available in 55 patients. Only four patients (7.2%) had PWV values above the 90th percentile and were classified as having increased arterial stiffness. All of these patients with arterial stiffness were normotensive and were not receiving antihypertensive therapy. The mean AIX@75 was 13% (Table 1).

Echocardiographic assessment showed a mean left ventricular mass of 29 g/m^2.7^, with a prevalence of LVH of 15.2% (95% CI: 4.8–25.6). When the cohort was stratified by LV mass z-score tertiles, no major clinical differences were observed across tertiles, except that patients in the highest tertile had higher BMI and CBP. Heart rate, PWV, and AIX@75 did not differ significantly between tertiles. As expected, increasing LV mass z-scores were strongly associated with greater PWTd and IVSTd (p < 0.001; Table 1).

To explore variables associated with cardiac remodeling, correlation analyses were performed using the structural components of LVH, namely PWTd and IVSTd. As shown in Figure 1, both PWTd and IVSTd were positively correlated with CBP and PWV. In contrast and unexpectedly, inverse associations were observed with AIX@75 and HR, although the latter did not reach statistical significance.

**Figure 1.**
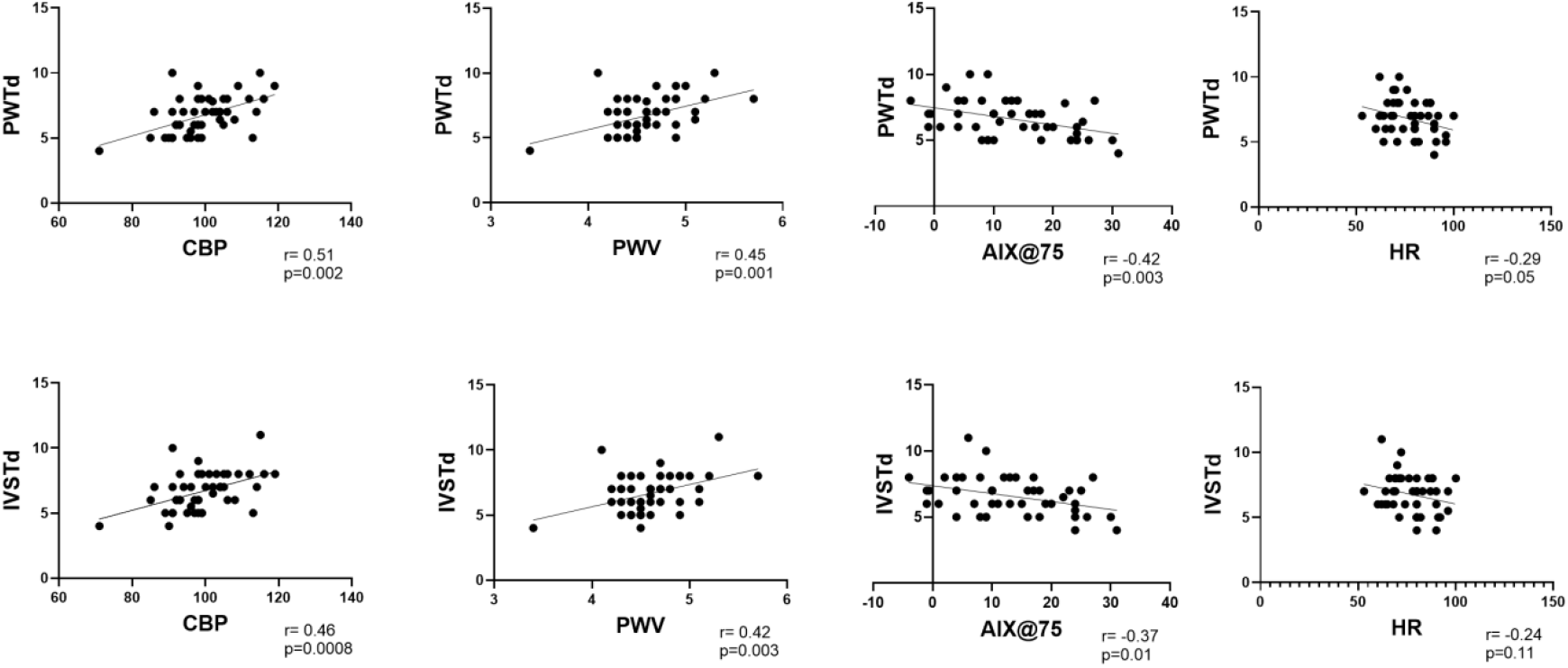
Correlations of Posterior Wall Thickness (PWTd) and Interventricular Septal Thickness (IVSTd). Posterior Wall Thickness (PWTd) and Interventricular Septal Thickness (IVSTd) exhibit significant positive correlations with central blood pressure (CBP) and pulse wave velocity (PWV) suggesting a link between increased arterial stiffness, central pressure and ventricular remodeling. In contrast, inverse associations were observed with the augmentation index (AIx@75) and heart rate (HR). The negative trend with HR is apparent but did not reach statistical significance.

To further evaluate the influence of central hemodynamics, blood pressure, and anthropometric variables on cardiac remodeling, simple and multivariable linear regression models were constructed using PWTd and IVSTd as dependent variables. Variables associated with PWTd are shown in Table 2A. As expected, CBP, peripheral blood pressure, and BMI were positively associated with PWTd. AIX@75 showed a negative association with PWTd, indicating that higher AIX@75 values were associated with thinner posterior walls. In multivariable analysis, AIX@75 remained inversely associated with PWTd after adjustment for central and peripheral blood pressure, age, and sex. This model explained up to 40% of the variability in PWTd (R^2^ = 0.40, p < 0.01). PWV was not independently associated with PWTd and was highly collinear with CBP; therefore, it was excluded from the final model.

**Table 2.** Variables associated with Posterior Wall Thickness (PWTd) including Augmentation_Index (2A) or Heart_Rate (2B)

| **Table 2A** | **Univariate $\beta$ (95% CI)** | **p-value** | **Multivariable $\beta$ (95% CI)** | **p-value** |
| --- | --- | --- | --- | --- |
| AIX@75 | -0.06 (-0.11, 0.02) | **<0.01** | -0.06 (-0.12, -0.002) | **0.04** |
| CBP | 0.08 (0.04, 0.12) | **<0.001** | 0.04 (-0.02, 0.11) | 0.18 |
| PAS24hr | 0.05 (0.02, 0.09) | **<0.01** | 0.02 (-0.02, 0.06) | 0.39 |
| BMI | 0.14 (0.05, 0.24) | **<0.01** | 0.10(0.004, 0.20) | **0.04** |
| Age | 0.11 (-0.03, 0.24) | 0.12 | -0.11(-0.25,0.03) | 0.21 |
| Sex (Male) | -- | -- | -0.16 (-1.15, 0.84) | 0.75 |
| **Table 2B** | **Univariate $\beta$ (95% CI)** | **p-value** | **Multivariable $\beta$ (95% CI)** | **p-value** |
| HR | -0.04 (-0.07, -0.001) | 0.05 | -0.04 (-0.08, -0.002) | **0.04** |
| CBP | 0.08 (0.04, 0.12) | **<0.001** | 0.03 (-0.03, 0.09) | 0.27 |
| PAS24hr | 0.05 (0.02, 0.09) | **<0.01** | 0.03 (-0.008, 0.07) | 0.12 |
| BMI | 0.14 (0.05, 0.24) | **<0.01** | 0.15 (0.05, 0.26) | **<0.01** |
| Age | 0.11 (-0.03, 0.24) | 0.12 | -0.09 (-0.24, 0.06) | 0.22 |
| Sex (Male) | -- | -- | 0.37 (-0.46, 1.19) | 0.37 |

Model 2A: $R^2=0.40$ ( $P<0.01$ ) MSE 1.14; Model 2b: $R^2=0.43$ ( $P<0.01$ ) MSE 1.12.

Given the inverse trend between HR and PWTd observed in Figure 1, the negative association between AIX@75 and PWTd in multivariable analysis, and the known dependence of AIX@75 on heart rate, an alternative model was constructed replacing AIX@75 with heart rate (Table 2B). This model showed improved performance (R^2^ = 0.44, p < 0.01) and lower error (MSE = 1.12). Heart rate remained independently and inversely associated with PWTd, indicating that lower HR was associated with greater posterior wall thickness. AIX@75 and heart rate were not included simultaneously in the same model because AIX@75 is mathematically adjusted for HR, precluding their treatment as independent variables.

Heart rate was also inversely associated with IVSTd after adjustment for BMI and central and peripheral blood pressure (Table 3B). In contrast, AIX@75 did not remain independently associated with IVSTd after multivariable adjustment (Table 3A).

**Table 3.** Variables associated with Interventricular Septum Thickness (IVSTd) including Augmentation_Index (3A) or Heart_Rate (3B)

| **Table 3A** | **Univariate $\beta$ (95% CI)** | **p-value** | **Multivariable $\beta$ (95% CI)** | **p-value** |
| --- | --- | --- | --- | --- |
| AIX@75 | -0.06 (-0.11, 0.01) | **0.01** | -0.05 (-0.12, -0.01) | 0.11 |
| CBP | 0.08 (0.03, 0.12) | **<0.001** | 0.04 (-0.04, 0.11) | 0.29 |
| PAS24hr | 0.05 (0.01, 0.08) | **0.01** | 0.02 (-0.02, 0.06) | 0.34 |
| BMI | 0.14 (0.05, 0.24) | **<0.01** | 0.13(0.02, 0.24) | **0.02** |
| Age | 0.11 (-0.03, 0.24) | 0.08 | -0.07(-0.24,0.10) | 0.42 |
| Sex (Male) | -- | -- | -0.10 (-1.21, 1.01) | 0.85 |
| **Table 3B** | **Univariate $\beta$ (95% CI)** | **p-value** | **Multivariable $\beta$ (95% CI)** | **p-value** |
| HR | -0.03 (-0.07, 0.01) | 0.14 | -0.04 (-0.09, -0.001) | **0.04** |
| CBP | 0.08 (0.03, 0.12) | **<0.001** | 0.03 (-0.04, 0.09) | 0.4 |
| PAS24hr | 0.05 (0.01, 0.08) | **0.01** | 0.02 (-0.01, 0.06) | 0.21 |
| BMI | 0.14 (0.05, 0.24) | **<0.01** | 0.17 (0.06, 0.29) | **<0.01** |
| Age | 0.11 (-0.03, 0.24) | 0.08 | -0.08 (-0.24, 0.08) | 0.33 |
| Sex (Male) | -- | -- | 0.29 (-0.6, 1.18) | 0.5 |

Model 3A: $R^2=0.38$ ( $p<0.01$ ) MSE 1.27; Model 3B: $R^2=0.40$ ( $P<0.01$ ) MSE 1.21

We performed a computational simulation to assess the impact of HR on PWTd. A uniform 10% increase in HR resulted in a mean reduction in PWTd of 0.36 mm and a marked decrease in the prevalence of posterior wall hypertrophy from 30.9% to 5.1% (p < 0.01) (Figure 2). In a simple logistic regression analysis, a 10% increase in HR was associated with a significant reduction in posterior wall hypertrophy (OR = 0.10; 95% CI: 0.01–0.44).

**Figure 2.**
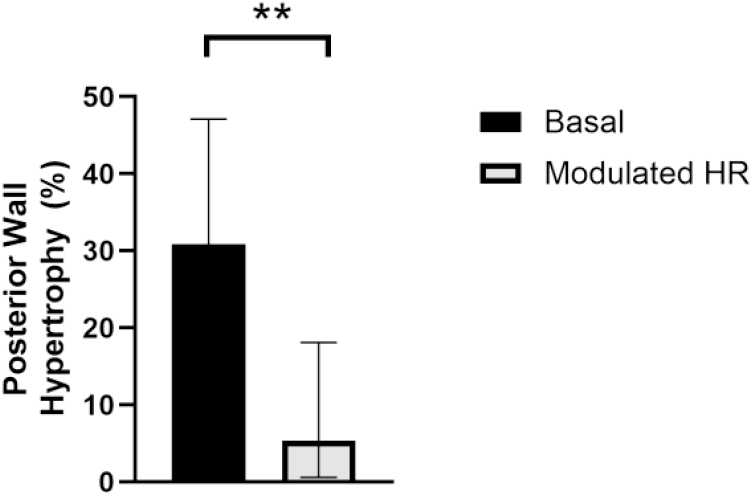
Effects of Heart Rate Modulation on Left Ventricular Remodeling. A computational simulation predicts that a 10% increase in heart rate, while keeping all other variables constant, reduces the posterior wall thickness diameter (PWTd) by 0.36 mm, leading to a decrease in prevalence from 30.1% (95% CI: 17.6-47.1) to 5.4% (95% CI: 0.6-18.1) (p<0.001).

Figure 3 illustrates representative pulse wave analyses from two patients with low and high heart rates. In the patient with lower HR, early wave reflection resulted in fusion of the forward (P1) and reflected (P2) waves, blunting the typical waveform morphology. In contrast, in the patient with an HR of 82 bpm, clear separation between P1 and P2 was observed, resulting in a higher AIX@75. These findings suggest that higher HR and the associated shortening of ejection duration shift the reflected wave later in the cardiac cycle, closer to diastole, thereby reducing early systolic load on the left ventricle. Conversely, at lower heart rates, prolonged systole favors early coupling of the reflected wave with the forward wave, increasing systolic ventricular loading.

**Figure 3.**
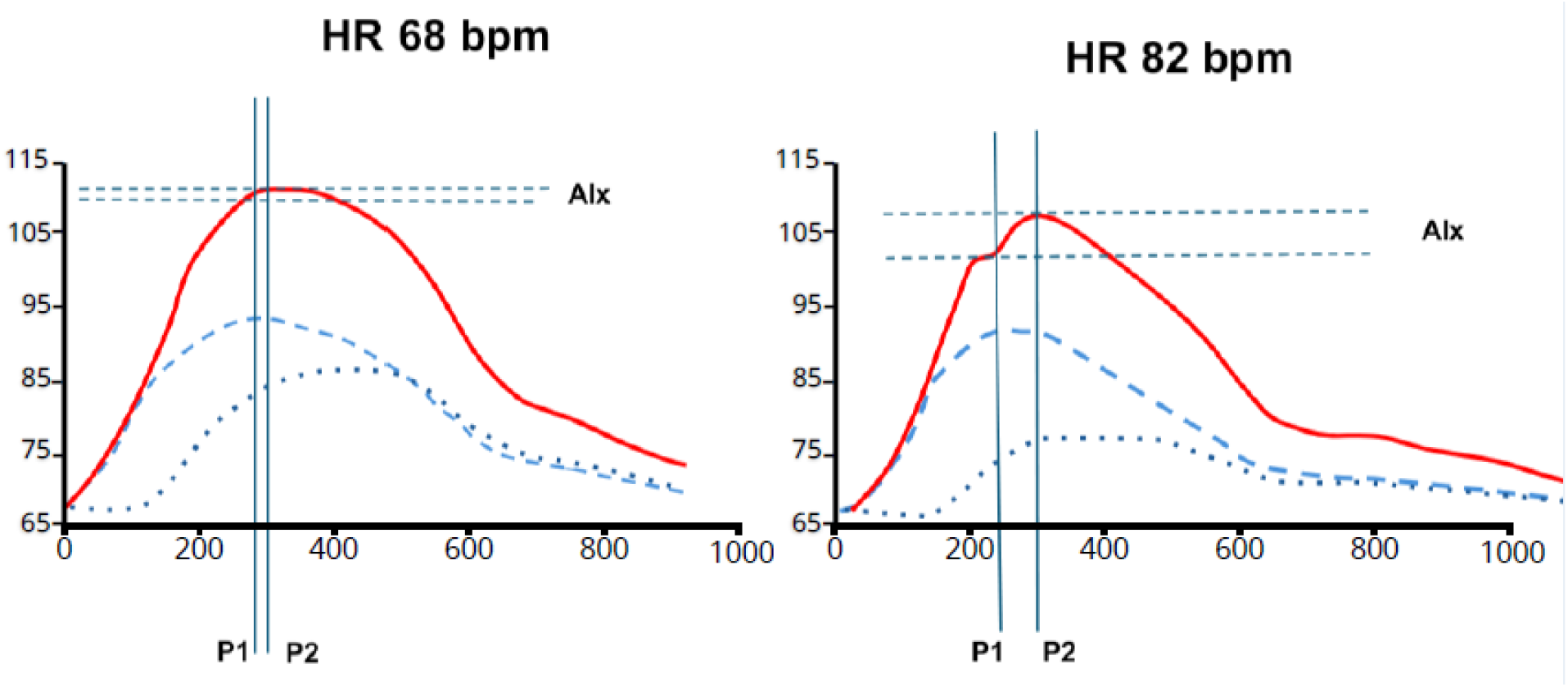
Representative pulse wave analysis in two patients with different heart rates. The arterial waveform (red) is composed of two main components: the forward wave (dashed line) and the reflected wave (dotted line). In the patient with a lower heart rate (68 bpm), the forward (P1) and reflected (P2) waves merge, blunting the typical waveform. In contrast, in the patient with a heart rate of 82 bpm, P1 and P2 are clearly separated, resulting in a higher AIx@75. These findings suggest that a higher heart rate shortens systole, shifting the reflected wave toward diastole and reducing early systolic load. Conversely, a lower heart rate prolongs systole, promoting earlier wave overlap and increasing left ventricular load.

## Discussion

In this study, we found that patients with long-term follow-up after CoAo repair exhibit LV remodeling, even in the absence of central hypertension. This remodeling was associated with lower HR and reduced AIX@75, consistent with an earlier return of the reflected pressure wave during systole. Importantly, a modeling simulation demonstrated that a modest increase in HR substantially attenuated LV remodeling.

A significant proportion of patients with repaired CoAo develop LV remodeling within a few years of intervention despite adequate blood pressure control. This phenomenon may contribute to early cardiovascular events and reduced long-term survival.(6) LV mass is influenced by pressure load, body size, and the mechanical properties of the aorta.(8,23,24) In this population, aortic characteristics may be persistently altered due to surgical scarring and vascular remodeling, creating conditions for abnormal and premature returning reflected waves.(8,10,11)

In our cohort, up to 14% of patients with repaired CoAo and non-significant residual gradient met criteria for LVH. However, a larger proportion exhibited subtle geometric changes that did not meet the 90th-percentile threshold, highlighting the continuous nature of LV remodeling.

Notably, most patients with LVH had normal peripheral and central blood pressure, suggesting that factors beyond pressure load contribute to myocardial remodeling in this setting. Similar observations have been reported by previous studies, which identified functional, but not structural, alterations during shorter follow-up periods.(10) The principal structural components of LV remodeling are posterior wall thickness and interventricular septal thickness.(20) We therefore modeled these parameters as linear outcomes, allowing independent evaluation of each component. In contrast, the calculated LV mass is derived from a cubic, nonlinear formula, which violates linear regression assumptions and may obscure component-specific effects.

In univariate analyses, we observed an inverse association between AIX@75 and both posterior wall and septal thickness. This finding contrasts with observations in the general population, in which higher AIX@75 is typically associated with LVH, owing to increased central systolic pressure resulting from wave reflection.(14,25) In our study, however, the effect of wave reflection appeared to be independent of blood pressure and instead related to the timing of the reflected wave within the cardiac cycle.

At lower HR, systolic duration is prolonged, increasing the likelihood that the reflected wave returns during early systole. In this scenario, the reflected wave merges with the forward wave (P1), resulting in minimal augmentation despite increased systolic load. This fusion may explain the paradoxical association of lower AIX@75 with greater LV remodeling without changes in CBP in CoAo. These observations prompted further evaluation of HR as an independent determinant of LV remodeling in this population.

Consistent with this hypothesis, lower HR was independently associated with greater LV remodeling, while simulated increases in HR reduced remodeling indices. Mechanistically, a higher HR shortens systole, thereby delaying the arrival of the reflected wave relative to the cardiac cycle, placing it closer to late systole or diastole. This temporal shift separates the reflected wave (P2) from the forward wave (P1), increasing AIX@75 while reducing early systolic load. Importantly, this rightward shift of wave reflection alters ventricular loading conditions without increasing absolute CBP, as illustrated in the exemplar patient waveforms (Figure 3).

Contrary to our hypothesis, other vascular parameters, such as PWV, a surrogate of arterial stiffness, were not independently associated with LV remodeling. Given their strong dependence on blood pressure and the presence of multicollinearity with AIX@75, these variables were not included in the final multivariable models.

Interestingly, the clinical phenotype of patients in the highest tertile of LV mass, or even those meeting criteria for LVH, did not show noticeable differences in resting heart rate or AIX@75 on unadjusted comparisons. Only after multivariable adjustment did the independent contribution of HR become evident. This observation underscores that LVM reflects the integrated effects of multiple interacting determinants, some of which may not be clinically apparent when considered in isolation. For example, a patient with a lower heart rate but slightly lower central blood pressure or normal body mass index may exhibit LV mass similar to that of another patient with a higher heart rate and modestly higher central blood pressure or body mass index, thereby masking the physiologic influence of HR on ventricular remodeling. Importantly, LV remodeling is a continuous process, and many patients do not reach the categorical threshold for LVH. Consequently, in our study, preliminary binary comparisons based on LVH status and heart rate did not detect meaningful associations, consistent with findings from a previous study.(26)

Ambulatory hypertension was present in 42% of the cohort; however, none of these patients demonstrated elevated CBP, suggesting a prominent role of peripheral pressure amplification in this population. These findings question the adequacy of treatment decisions based solely on peripheral blood pressure measurements in patients with repaired CoAo and support the need for systematic assessment of central hemodynamics. Future studies should evaluate the relative prognostic significance of peripheral versus central blood pressure in this population.

Among patients with ambulatory hypertension, treatment rates were relatively high (approximately 80%) in this tertiary center, yet blood pressure control was achieved in only 58%. Notably, about 10% of patients with repaired CoAo were receiving beta-blockers. In light of our findings, the routine use of beta-blockers in this population, in the absence of other compelling indications, warrants reconsideration. Prospective studies are needed to clarify the impact of heart rate-lowering therapies on ventricular remodeling and long-term cardiovascular outcomes in repaired CoAo. Conversely, antihypertensive therapies that do not reduce heart rate, or that modestly increase HR, such as direct vasodilators, may theoretically shift wave reflection later in the cardiac cycle and attenuate adverse ventricular remodeling.

Importantly, these considerations may extend beyond the management of hypertension. In patients with repaired CoAo who are normotensive but exhibit LV hypertrophy or early remodeling, modulation of heart rate may represent a novel therapeutic target independent of blood pressure reduction. Whether modest increases in HR can favorably alter ventricular loading conditions and structural remodeling in this population requires prospective evaluation. Carefully designed interventional studies will be necessary to determine the safety, feasibility, and long-term cardiovascular impact of heart rate– modulating strategies in patients with repaired CoAo.

This study has several limitations. First, its cross-sectional design precludes causal inference, and all observed relationships should be interpreted as associations. Prospective longitudinal studies are needed to confirm these findings and determine whether heart rate modulation can reduce left ventricular remodeling and long-term cardiovascular risk in patients with repaired CoAo. Ultimately, interventional trials will be required to establish clinical utility. Second, the present findings apply specifically to patients with repaired CoAo or hemodynamically non-significant residual coarctation. It remains unclear whether these observations can be extrapolated to patients with unrepaired CoAo, particularly given recent in silico models suggesting that lower heart rate may reduce wall shear stress and exert favorable hemodynamic effects in that setting.(27) Third, the retrospective nature of the study may introduce bias and unmeasured confounding. Certain variables, such as interobserver variability in echocardiographic measurements and incomplete data capture, could not be fully controlled and may have contributed to systematic error. Fourth, HR was assessed at a single time point and may not reflect long-term autonomic tone or circadian variability, potentially leading to misclassification of the true exposure of interest. Fifth, this study was conducted at a single tertiary care center, which may limit the generalizability of the findings to other CoAo populations with different clinical complexity, surgical techniques, or follow-up strategies. Finally, vascular mechanics were assessed using an oscillometric device that estimates forward (P1) and reflected (P2) pressure waves rather than direct measurements obtained by applanation tonometry. This approach was chosen because of its practicality, shorter acquisition time, suitability for pediatric patients, and the availability of age- and sex-specific reference values. Nevertheless, these measures are model-based estimates rather than direct recordings and may introduce systematic measurement error.

## Conclusion

In patients with repaired CoAo, LV remodeling persists despite normal peripheral and central blood pressures and is independently and inversely associated with HR and indices of wave reflection timing. These findings suggest that ventricular remodeling in this population is influenced not only by pressure load but also by the temporal interaction between cardiac cycle duration and arterial wave reflections. Modulation of HR may therefore represent a novel, blood pressure–independent target to mitigate adverse ventricular remodeling. Prospective studies are needed to confirm these observations and to determine whether heart rate–guided therapeutic strategies can improve long-term cardiovascular outcomes in patients with repaired CoAo.

## Data Availability

All data produced in the present study are available upon reasonale request to the authors.

## List of abbreviations

CoAo: coarctation of the aorta
LVH: left ventricular hypertrophy
ABPM: ambulatory blood pressure
PWTd: posterior wall thickness in diastole
IVSTd: interventricular septal thickness in diastole
AIX@75: heart rate–adjusted augmentation index LV: left ventricular
HR: heart rate
BMI: body mass index
LVM: left ventricular mass
PWV: pulse wave velocity
CBP: central blood pressure
P1: forward wave
P2: reflected wave

## Declarations

### Ethics approval and consent to participate

The current study was approved by the Ethics Committee of the Government of the Autonomous City of Buenos Aires, Hospital de Niños “Dr. Ricardo Gutiérrez” (approval code: 16417). Informed consent was waived due to the retrospective nature of the study.

### Consent for publication

All authors have read the manuscript and approve it for publication.

### Availability of data and materials

The datasets generated and analyzed during the current study are available from the principal investigator on reasonable request.

### Authors’ contributions

MV contributed to data collection, study design, data analysis, and overall structuring of the manuscript. LEM, CGM, AS, MR: Participated in data collection and conducted thorough reviews of the manuscript. CAR contributed to the study design, data analysis, and manuscript generation.

## Acknowledgements

The authors thank Cooperadora de Acción Social (COAS), an Argentinian non-profit foundation, for providing central blood pressure devices.

